# Managing the evidence infodemic: Automation approaches used for developing NICE COVID-19 living guidelines

**DOI:** 10.1101/2022.06.13.22276242

**Authors:** Mariam Sood, Steve Sharp, Emma McFarlane, Robert Willans, Kathryn Hopkins, Justine Karpusheff, Fiona Glen

**Affiliations:** National Institute for Health and Care Excellence 2^nd^ Floor, 2 Redman Place London E20 1JQ; National Institute for Health and Care Excellence Level 1A, City Tower, Piccadilly Plaza Manchester M1 4BT

**Keywords:** Automation, Surveillance, Guidelines, COVID-19, Evidence management, Literature screening, Clinical Trials, machine learning classifier, pattern matching, regular expressions

## Abstract

**Background and Objectives:** The National Institute for Health and Care Excellence (NICE) produces evidence-based guidance and advice for health, public health and social care practitioners in England and Wales. Between March 2020 and March 2022, NICE produced 24 COVID-19 guidelines to support healthcare workers during the COVID-19 pandemic. This article outlines three automation strategies NICE utilised to facilitate faster processing of evidence on COVID-19 and describes the value of those approaches when there is an increasing volume of evidence and demand on resources.

**Study Design and Setting:** Text classification using machine learning, and regular expression-based pattern matching were used to automate screening of literature search results. Relevant clinical trials were tracked by automated monitoring of clinical trial databases and Pubmed.

**Results:** The strategies discussed here brought considerable efficiencies in the processing time without impacting on quality compared to equivalent manual efforts. Additionally, the paper illustrates how to incorporate automation into established processes of the evidence management pipeline.

**Conclusions:** We have demonstrated through testing and use in live guideline development and surveillance that these are effective and low risk approaches at managing high volumes of evidence.

**Highlights:**

- To illustrate how NICE utilised automation to handle the Covid-19 ‘infodemic’-managing the ‘infodemic’ of evidence surveillance is a shared global issue.
- To outline automation strategies to facilitate faster processing of evidence, especially when there is an increasing volume of evidence and demand on resources.
- How automation can be included in established processes without disrupting business as usual operations.
- Automation can take many forms, and depending on risk appetite, can be supplemented with manual checking.
- Automation can be adopted easily with the right tools and techniques.

## 1. Introduction

Between March 2020 and March 2022, NICE produced 24 COVID-19 guidelines to support healthcare workers during the COVID-19 pandemic [1]. Throughout the pandemic, it’s been essential to continuously monitor the rapidly evolving evidence base on COVID-19 through a frequent broad search, to ensure the recommended advice remains current. Key ongoing trials are monitored for publications so studies can be considered for impact on guideline recommendations as soon as published. However, it can be challenging to keep abreast of trial readouts in a topical area such as COVID-19.

A weekly broad search covers all major subscription databases, freely accessible databases and pre-print servers. For a topical subject such as COVID-19, the weekly search generates approximately 3000 records. The weekly screening of the search results is a significant task, which when conducted manually took one systematic reviewer 2 to 3 days. Since the start of 2021, we implemented several automation approaches which provided considerable efficiencies in retrieving key evidence more quickly and enabling quicker identification of evidence from completed trials.

The focus of this paper is to discuss the automation approaches we employed for COVID-19 surveillance at NICE, which are techniques drawn from the wider field of natural language processing, a sub-field of AI dealing with text processing. Specifically, we talk about three automation projects which addressed various aspects of our surveillance evidence pipeline:

1. Custom trained machine learning classifiers to automate study screening,
2. Rule based pattern matching to automate categorising studies to sub-topics relevant to NICE COVID-19 guidelines and
3. Automated monitoring of the progress of ongoing clinical trials.

The first two approaches are directly relevant to screening of literature search results, while the third approach automatically tracks changes in the status of clinical trials. Another main contribution of this paper is to illustrate how automation can be incorporated into guideline surveillance when there are established standard processes that are performed manually. The strategies outlined here facilitate faster processing of evidence, especially when there is an increasing volume of evidence and demand on resources.

## 2. COVID-19 Search and Surveillance at NICE

NICE surveillance includes monitoring of key ongoing studies, in addition to searching for relevant published evidence. Currently there are more than hundred COVID-19 trials being tracked by NICE surveillance. However, monthly checking for changes in trial status is resource intensive, taking around 2 days manual effort.

The COVID-19 search process began with a daily and subsequently weekly broad search covering all major subscription databases, freely accessible databases and pre-print servers. Each week, a systematic reviewer performed initial ‘triage’ screening on new evidence and categorised evidence into sub-topics of interest. Evidence was excluded if it was of low relevance to the guidelines or ineligible against review protocols. Using predetermined criteria, reviewers conducted second level screening and determined whether the evidence fell into: impactful evidence, supporting evidence or potential future impact, and relevant actions followed. To date, the number of abstracts in the NICE’s COVID-19 surveillance database stands at 300,000, of which around 18,850 have been screened as possible ‘includes’ (deemed relevant to NICE guidelines). This suggests that the majority of the search results were irrelevant for our purposes. However, low precision is a common phenomenon in broad population searches where there is a need to maximise recall of relevant studies. Managing this volume of evidence on a weekly basis is a resource intensive approach that is amenable to automation.

## 3. Approach 1: Using machine learning classifiers to automate study screening

### 3.1 The approach

The first approach involves using a custom trained machine learning text classifier to identify the search results as potential includes or definite excludes. Since a large proportion of the weekly search results were excludes, this approach had the potential to provide considerable noise reduction without removing an unacceptable proportion of relevant studies.

The machine learning classifier building tool that is implemented as part of the EPPI-4 reviewer software [2] was used to train a custom COVID-19 classifier that distinguished between relevant and irrelevant studies in the surveillance search. The EPPI-4 machine learning classifier building tool (hereafter called the EPPI-4 custom classifier) implements a logistic regression model that utilises a “bag-of-words” approach using tri-grams without word stemming [3].

The EPPI-4 custom classifier was trained and tested in three cycles, using the cumulative search results and screening decisions from March 2020 to December 2021.

The trained and tested classifier (COVID classifier) from each cycle was utilised to automate the screening decisions within the COVID-19 workstream at NICE. Figure 1 illustrates the screening process with COVID classifier included.

**Figure 1:**
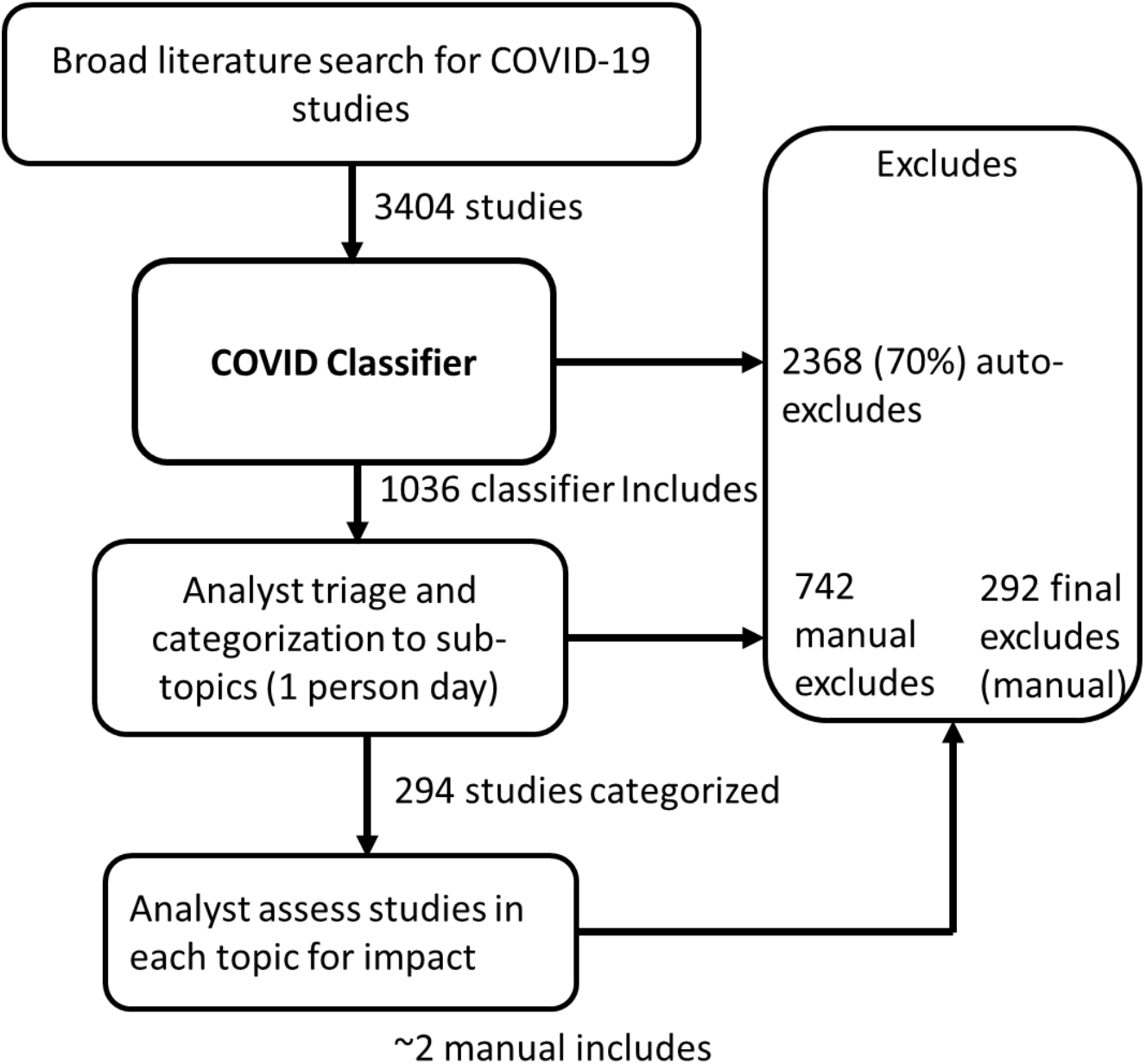
Surveillance screening with trained machine learning classifier, representative sample from week of 10^th^ February 2022

### 3.2 Methods

The EPPI-4 custom classifier was trained using the COVID-19 surveillance data and the associated screening decisions. The trained classifier was then applied to ‘unseen’ studies to output a confidence score for each study indicating how relevant it was for NICE COVID-19 surveillance. The confidence score is on a continuous scale rather than a binary decision of relevance. This score can then be converted into a binary decision by applying a ‘threshold’, below which the citation records will not be screened manually. Any studies which fall below the threshold are excluded automatically, saving manual screening time. Before adopting the trained classifier, it was tested on a separate hold-out dataset with known screening decisions to ensure the performance was reliable and within the accepted risk levels. The accuracy and recall at various threshold settings were evaluated on the hold-out dataset, and the threshold was chosen which provided greater than 96% recall. Precision was calculated but for the COVID-19 search this is not as critical a measure of classifier effectiveness as reduction in search output. This is because precision will always be low in broad population searches where there is a very low yield of includes relative to the size of the search output. The reduction in search output is a more valuable indicator of time saving efficiency, together with the percentage of false negatives to assess the impact on recall.

Additional testing was carried out prospectively for 4 weeks on live search outputs, where the studies automatically excluded by the classifier were manually screened, and any false negative excludes noted. This precautionary approach was conducted to mitigate the risk of false negative studies which were relevant but erroneously excluded. The data were then reviewed, and a decision made on whether to rely on the classifier without manual checking of the excludes, at the threshold relevance score.

The COVID classifier has been trained three times during the course of the pandemic to ensure the classifier adapts to the dynamic changes in the COVID scientific literature. The number of studies that were included in each training and test cycles are as shown in Table 1.

**Table 1:** Training and Tests datasets used in three cycles of classifier builds

| Training cycles | Training data |  | Test data |  |
| --- | --- | --- | --- | --- |
|  | Includes (n) | Excludes (n) | Includes (n) | Excludes (n) |
| Cycle1<br>(03/2020-05/2020) | 2724 | 41,176 | 1244 | 13,822 |
| Cycle2<br>(03/2020 - 03/2021) | 18,572 | 147,374 | 890 | 20,791 |
| Cycle3<br>(03/2020 - 11/2021) | 22,868 | 240,453 | 1251 | 17,370 |

### 3.3 Results

After each training cycle, the classifier threshold was chosen to ensure the false negatives are within the acceptable risk level, while still providing adequate noise reduction. The metrics used for deciding the threshold are summarised in Tables 2-4. Based on the results, a threshold of 30% relevance score was chosen after cycle 1 training. At this threshold nearly 30% of the total test dataset were automatically excluded, with 5.6% of false negatives (at 1^st^ level screening decision). The threshold was raised to 60% after cycle 2 training because at this threshold the retrained model automatically excluded 82.8% of the total test dataset with 22.6% of false negatives (1^st^ level screening decision), none of which were impactful studies and prospective sample checking of the excludes at this threshold indicated minimal risk of missing impactful studies. After cycle 3 training, the threshold was reduced to 40% because the model performance was comparable to cycle 2 at this threshold (1.2% larger reduction in database size, 4.7% increase in false negatives) but at higher thresholds was less sensitive with significantly higher false negatives. Further precautionary testing of excludes at the 40% threshold indicated negligible risk of missing impactful studies.

**Table 2:**
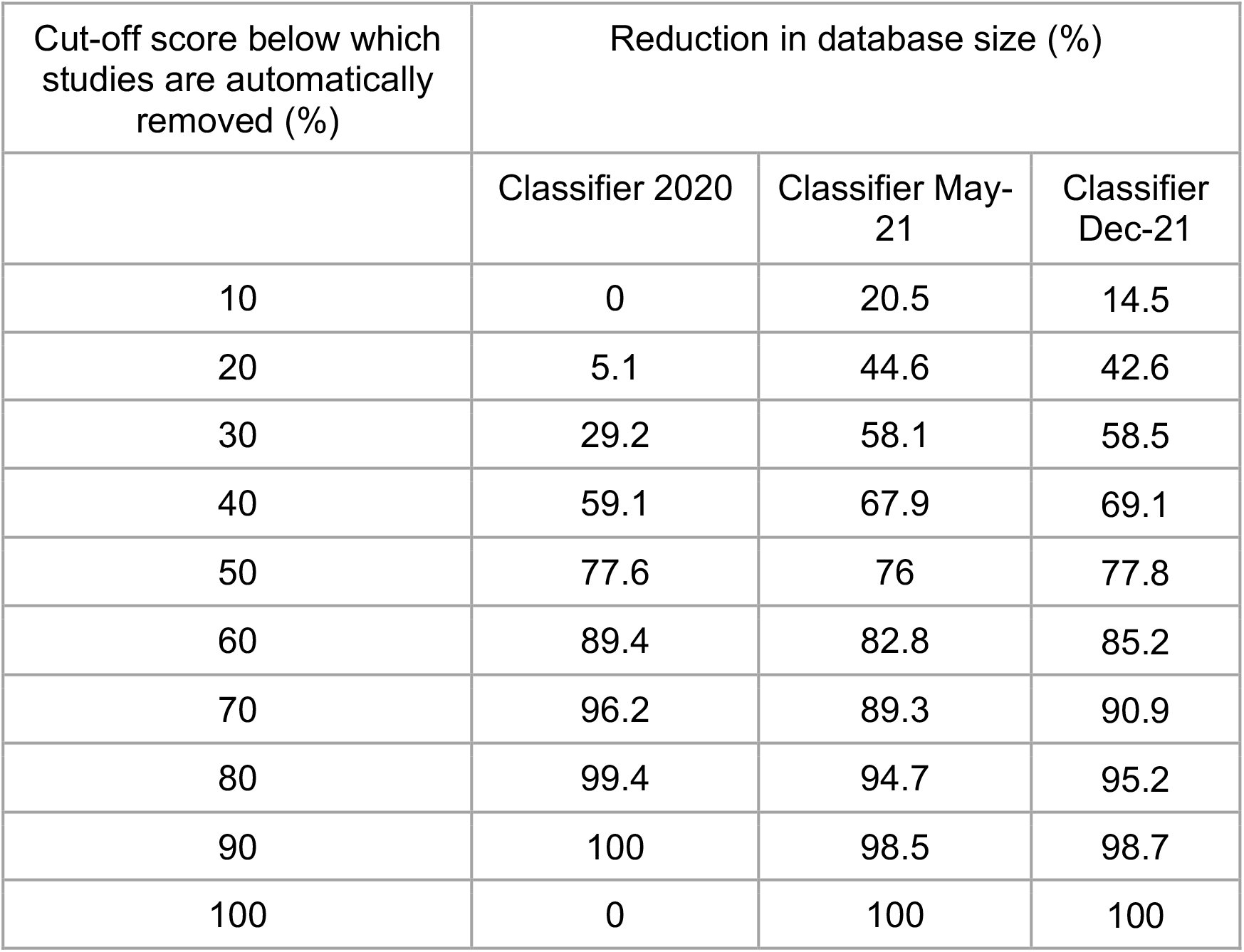
Classifier performance: reduction in database size at various thresholds

**Table 3:** Classifier performance: % false negatives at various thresholds

| Cut-off score below which studies are automatically removed | % triage false negatives |  |  |
| --- | --- | --- | --- |
|  | Classifier May 2020 | Classifier May-2021 | Classifier Dec-2021 |
| 10 | 0 | 0 | 0.4 |
| 20 | 0.1 | 0.3 | 3.6 |
| 30 | 5.6 | 1 | 5.5 |
| 40 | 25 | 4.3 | 9 |
| 50 | 49.9 | 10.9 | 21.7 |
| 60 | 74.5 | 22.6 | 42.4 |
| 70 | 90.9 | 40.8 | 64 |
| 80 | 99.2 | 62.5 | 81.1 |
| 90 | 100 | 85.5 | 95.2 |
| 100 | 0 | 100 | 100 |

**Table 4:** Precision and Recall at various thresholds for each cycle of classifier builds based on 1^st^ level triage includes. All values are in percentage.

|  | Cycle 1 |  |  | Cycle 2 |  |  | Cycle 3 |  |  |
| --- | --- | --- | --- | --- | --- | --- | --- | --- | --- |
| Threshold | 30 | 40 | 60 | 30 | 40 | 60 | 30 | 40 | 60 |
| Precision | 11 | 20.4 | 29.8 | 9.7 | 12.2 | 18.5 | 15.3 | 19.8 | 26.2 |
| Recall | 96.9 | 84.4 | 32.3 | 99.0 | 95.7 | 77.4 | 94.5 | 91 | 57.6 |

### 3.4 Discussion

The results above show that custom-built classifiers can significantly reduce the manual sifting burden without compromising recall of high impact studies, such as randomised controlled trials and systematic reviews, albeit with a lower but tolerable recall of lower impact studies. They are particularly useful for clearly defined searches, such as in the NICE COVID-19 portfolio of multiple distinct guidelines which share common core population terms, but where the search results contain considerable noise that is laborious to reduce using conventional methods. The algorithms can also automatically learn from the data, and hence can be more sustainable when there are dynamic changes in the dataset, such as for COVID-19.

## 4 Approach 2: Rule based pattern matching to automate categorising studies to sub-topics

### 4.1 The approach

In approach 2, we discuss an NLP software (hereafter called ‘categorisation software’) developed in-house to automate the triage sifting stage of manual surveillance, where the analyst determines whether each study is relevant to the guidelines and categorises it further into sub-topics of interest for more in-depth assessment. The technique utilized is rule based pattern matching using regular expressions and is a powerful technique to codify manual screening when there are discernible patterns in the text.

The categorisation software takes a set of studies (titles and/or abstracts) as input. For each study, the software determines whether the study belongs to any of the 35 implemented categories. It is possible for a single study to be relevant to multiple topic areas. In addition, the software identifies whether the study is an RCT (randomized control trial), a systematic review/meta-analysis, or an observational study (case-control, cross-sectional, cohort). The software produces 3 outputs: 1) the list of studies it received as input, where each study in the list is tagged with the topic categories and study design it falls under, 2) visual summary of the distribution of 35 topic areas in the list of studies, i.e., frequency graph of 35 topics in the list of studies and 3) visual summary of the distribution of study designs in the list of studies. Output 1 is exported as an Excel spreadsheet. Outputs 2 and 3 are as depicted in Figure 2.

**Figure 2:**
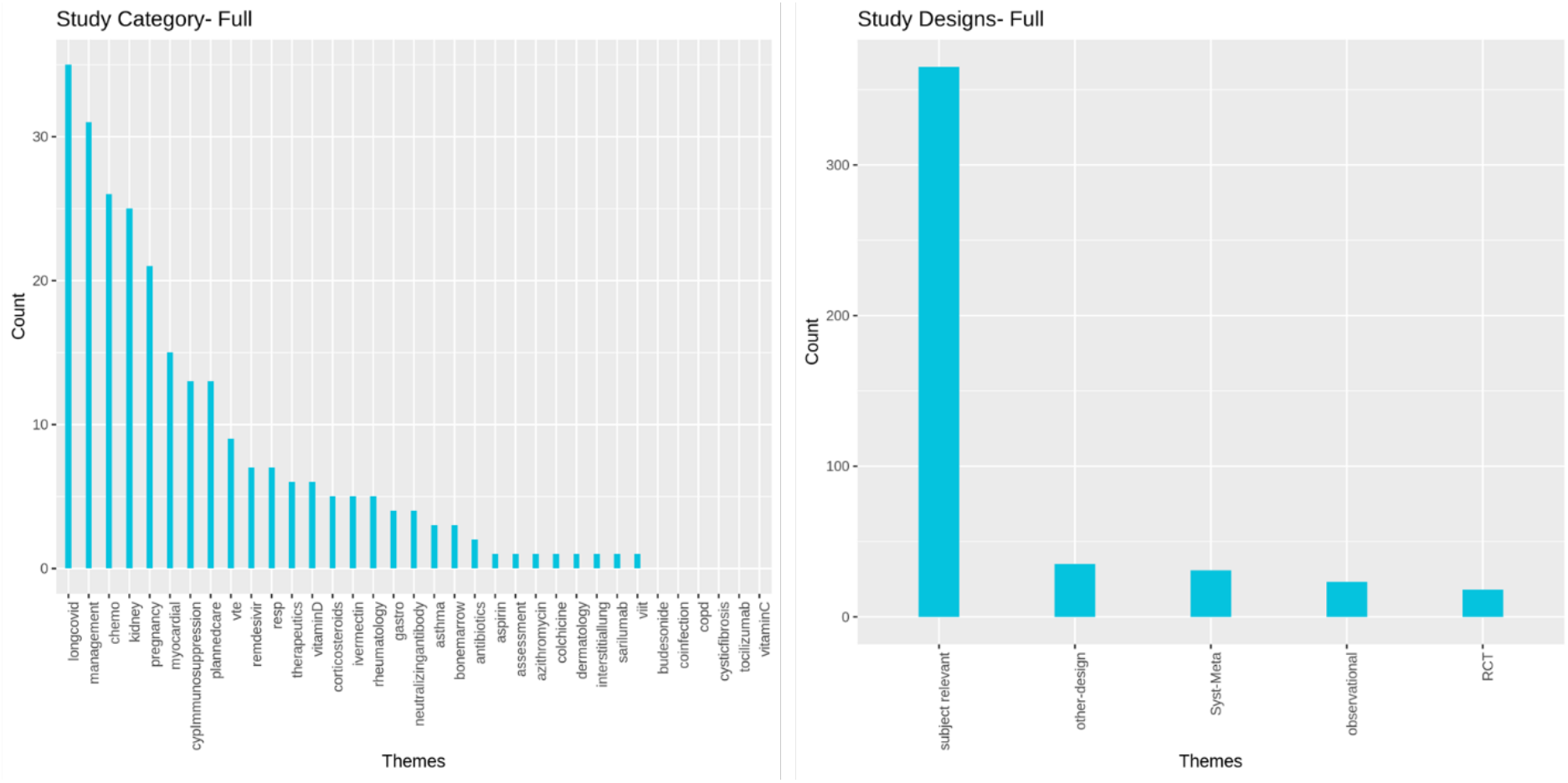
Visual summary outputs from Automated categorization of COVID studies software

The stages where automated categorisation is utilized within COVID surveillance is shown in Figure 3. This module can be applied directly on the output of the search results or on the output of the classifier includes/ excludes.

**Figure 3:**
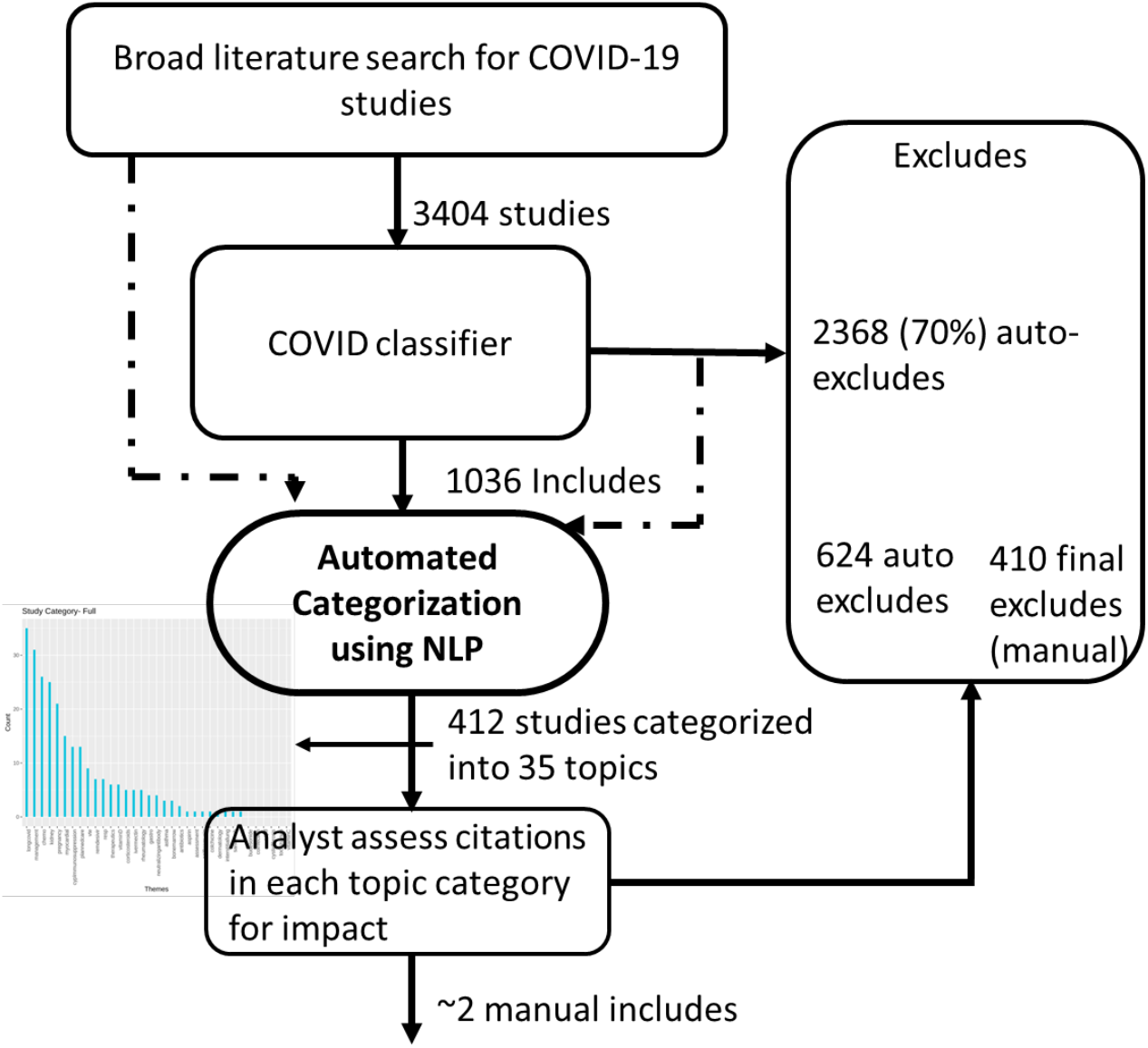
Surveillance screening with trained machine learning classifier and automated categorization, representative sample from week of 10^th^ February 2022

### 4.2 Methods

The categorisation software discussed here is custom developed to identify the COVID topic areas of interest. The software is implemented in Python [4], using Python’s NLP package spaCy [5]. The technique utilised is rule based pattern matching. The software implements 35 ‘pattern matchers’, where each ‘pattern matcher’ identifies the pattern corresponding to the respective topic of interest. In addition, there are three additional pattern matchers to identify the study designs (RCT, Systematic Review/Meta-analysis, observational design).

To simplify the potential variations of patterns that need to be identified, the titles/abstracts of each study are pre-processed. Two pre-processing steps are carried out. 1. change the text into lower case and 2. replace all punctuations with spaces to simplify the combinations of patterns to look for.

**Pre-processing Example**

Original title: Post-acute COVID-19 syndrome negatively impacts health and wellbeing despite less severe acute infection

Pre-processed title: post acute covid 19 syndrome negatively impacts health and wellbeing despite less severe acute infection

The text of titles/abstracts are searched for relevant patterns. The pattern matching code uses spaCy’s rule matching engine-Matcher [5], coupled with regular expressions where complex patterns need to be identified. The software was developed with the assistance of domain experts who provided subjective knowledge that govern the rules. Past screening done manually were also used as reference during development. The software was fine-tuned based on user testing during the month of November 2021.

**Pattern matcher rules based on regular expressions for identifying Vitamin D studies** [[{‘LOWER’: {‘REGEX’: (‘^(vitamin|vit|hydroxyvitamin|hypovitaminosis)$’)}}, {‘LOWER’: {‘IN’: [‘d’, ‘d3’]}}], [{‘LOWER’: {‘REGEX’: (‘^(cholecalciferol|ergocalciferol|calcifediol)$’)}}]]

The software was further tested and validated in two automated testing cycles. In cycle 1 testing, the triage includes for the months of April and May 2021 were used. In cycle 2 testing, the software was fine-tuned and further tested on triage includes for the months of September and October 2021.

The software is available at https://github.com/nice-digital/SciLiteratureProcessing

### 4.3 Results

The categorisation software was fine-tuned based on user testing and feedback during the month of Nov 2021. The software was further tested and validated in two automated testing cycles, using the first level triage includes data from April-May 2021 (868 studies) and Sept-Oct 2021 (934 studies). For the April-May 2021 triage includes, the software automatically excluded 358 studies as not belonging to any categories of interests. There were no impactful studies amidst the 358 studies. For the Sept-Oct 2021 triage includes, the software automatically excluded 177 studies without missing any studies with immediate impact. The cycle 1 and 2 testing are summarised in figures 4 and 5 respectively.

**Figure 4:**
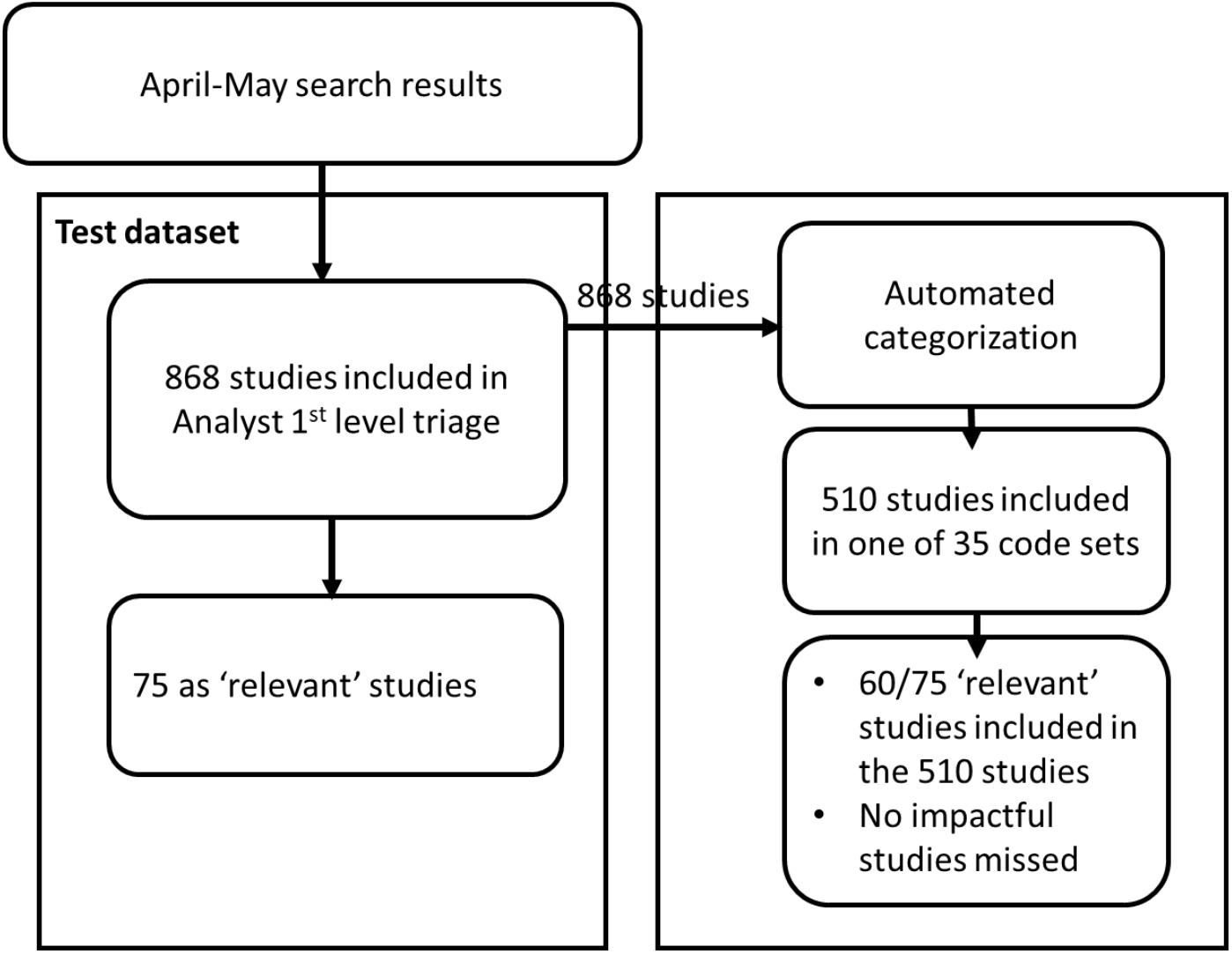
Cycle 1 Testing using April 2021 – May 2021 triage includes

**Figure 5:**
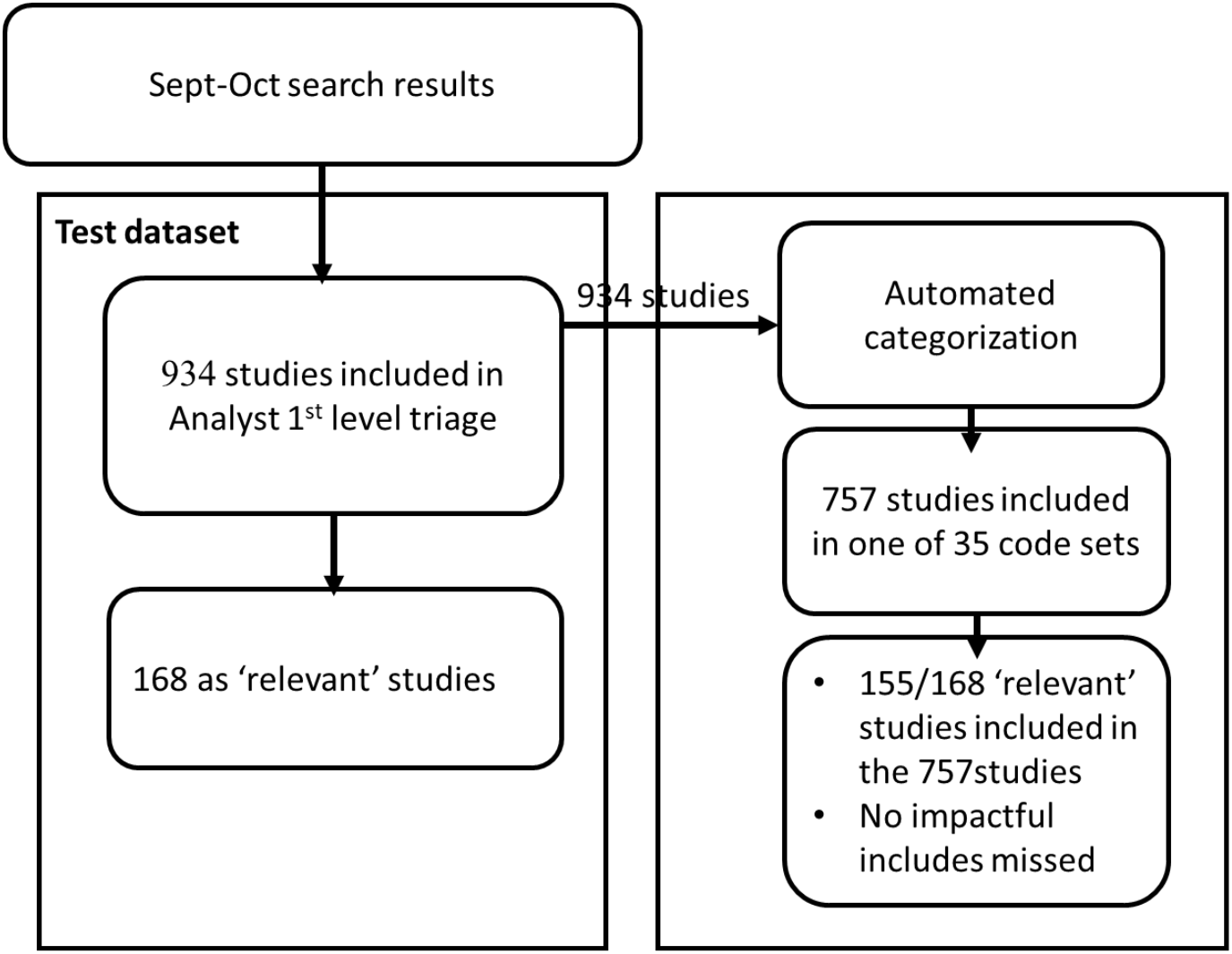
Cycle 2 testing using September-October 2021 triage

The software is designed to balance risk and noise. For e.g., for April-May 2021 dataset, while analysts included 868 records in their first level includes, software only included 510, which implies around 41% of the analyst includes were excluded automatically by the software. Of those excluded by the software, there were no impactful includes and 15 nice-to-have studies. This implies better noise reduction while keeping the risk low. Where studies are missed, they mainly fall into the noisy categories. The software is especially useful for categories which have well defined patterns that differentiate them. Majority of the categories amongst the 35 categories have well defined pattern rules that identify them.

### 4.4 Discussion

The automation technique used is ‘rule based pattern matching’ that implies the patterns to be identified are pre-specified in the software. Hence this method works well when we know what we are looking for. In an organisation such as NICE where systematic domain knowledge is employed in decision making, regular expressions provide a low-risk route to codify and automate the otherwise manual processes. The software is not an auto-learning method and can identify only patterns that it is programmed to identify. However, this technique does not require training, and works on datasets of any size.

## 5 Approach 3: Automated trial tracking

### 5.1 The approach

Thirdly, we describe two automated trial tracking approaches and compare those with the currently used standard manual approach for monitoring progress of ongoing trials. In the first automated approach (automated approach – registries), the trial identification number was used to query the database APIs, with the results of the query formatted into CSV files. For returning data from clinicaltrialsregister.eu, the ‘ctrdata’ R package [6] was used to return data. For ISRCTN [7], NIHR Journals Library [8] and ClinicalTrials.gov [9], direct calls to the API were made. In the second approach (automated approach – PubMed), a script was generated which queries the PubMed API [10] for the trial identification number. Results of the query were formatted into an Excel spreadsheet. The output provides a list of publications related to a trial identification number and the relevant URL. A daily email alert system was set up to generate notifications when there is a change of status of the trial, for example the trial status has changed from ongoing to completed.

### 5.2 Methods

To assess the performance, the two automated approaches were run alongside the manual approach for a 3-month period (July 2020 to September 2020). A convenience sample of ongoing trials identified by NICE surveillance was used for the test providing they were: 1) Ongoing at the time of the study, 2) Listed on one of the trial databases included in this study (ISCRTN, NIHR Journals Library, clinicaltrialsregister.eu and ClinicalTrials.gov).

For the manual approach, the current status of each record was established monthly by checking the related record on the trial databases or NIHR Journals Library repository and any change in status, or pertinent information, was recorded. Additionally, manual web-based searches were conducted on PubMed to identify a related publication associated with the trial. Records with a related publication were changed in status to closed events (i.e. removed from the tracking).

For both automated approaches, scripts were run daily. Any changes in a trial record flagged through the automated approach were checked for a relevant publication on the same day and the event closed if a publication was identified and the event recorded

The outputs of each approach were compared to determine which identified the most trials and in the quickest timeframe. To be able to calculate these measures, any publication identified by any of the strategies assessed was considered the gold standard.

The average time to event for each approach was determined by calculating the difference between the date the event was published online and the date the event was first identified by any approach.

To determine the resource impact of each option, the time taken to apply the approach was logged each month in hours by the reviewers. This data was used to calculate the average time taken (as a proxy for resource use) to identify one completed event in minutes.

### 5.3 Results

Over a 3-month period, the two automated approaches for tracking trials were compared with the standard manual approach.

Both automated approaches were feasible and had benefits over the manual approach, in summary:

- The time to event (defined as identification of a trial) was shortest for the automated approach – PubMed (mean 11 days) and longest for the manual approach (mean 108 days).
- The automated approach – PubMed was least resource intensive and quicker for a reviewer to identify a trial because any notification via this route was always related to a completed trial publication. For the other two approaches, further investigative work was required meaning a notification of change in trial status did not always result in identification of a publication. However, the automated approach – registries still reduced the time taken to identify a trial compared with the manual approach.
- Notably, the automated approach using PubMed identified more trials than the automated approach using trial registries, and was considered much easier technically to implement. The registries will always be limited by the different fields they include and how often researchers update records.

### 5.4 Discussion

The results of testing indicate it is feasible to automate trial tracking and deliver benefits in terms of time to trial identification and resource use. In particular, automating monitoring of ongoing trials may be a good strategy for ‘living’ guidelines where there is a focus on updating recommendations in response to emergent evidence soon after publication of results.

However, the automated approach is limited in terms of whether ongoing trials of interest are registered on trial databases and not all trial registries currently have developed APIs. Therefore, it is likely that an element of manual tracking will also need to be retained for certain studies and monitoring of non-trial events. The automated approach using PubMed performed better than the automated approach using trial registries and further work should be done to determine whether this should be the only automated approach considered going forward.

## 6. Conclusion

The three approaches discussed here present novel ways to automate guideline surveillance, reducing manual burden and expediting the throughput of evidence in the surveillance pipeline. There is often a hesitancy in using automation, and in particular machine learning based approaches due to the potential risk of missing key studies. We have demonstrated through testing and use in live guideline development and surveillance that these are effective and low risk approaches at managing high volumes of evidence on a regular basis.

## Data Availability

All data referred to in the manuscript are available upon reasonable request to the authors

https://github.com/nice-digital/SciLiteratureProcessing

## Abbreviations

NLP: Natural language processing
ML: Machine learning
API: Application programming interface

## Author contributions

Mariam Sood: Conceptualization, Methodology, Software implementation, Formal analysis of Approach 2; Writing-original draft preparation

Steve Sharp: Conceptualization, Methodology, Formal analysis of Approach 1; Writing-original draft preparation

Emma McFarlane: Conceptualization, Methodology, Formal analysis of Approach 3; Writing-original draft preparation.

Robert Willans: Conceptualization, Methodology, Software implementation of Approach 3; Writing-reviewing and editing.

Kathryn Hopkins: Conceptualization, Methodology and Validation of Approach 1; Writing-reviewing and editing.

Justine Karpusheff: Conceptualization, Project sponsor-Approach 1 and Approach 2; Writing-reviewing and editing

Fiona Glen: Conceptualization, Project sponsor-Approach 2; Writing-reviewing and editing.

## Acknowledgements

Thank you to Olivia Crane, Sarah Boyce, Toby Mercer, Omnia Bilal, Meena Tafazzoli, Rachel Archer, Maria Majeed and Ceri Williams for domain insights, assistance in data curation and validation utilized in Approaches 1 and 2.

Thank you to Monica Casey, Catherine Jacob and Niamh Knapton for testing Approach 3 and Juliana Sanabria for assistance with data analysis of Approach 3.

## Funding sources

None

## Declaration of interests

The authors declare no competing interests

## Notes

### Competing Interest Statement

The authors have declared no competing interest.

### Funding Statement

The authors received no specific funding for this work.

